# Prevention of post-concussion-like symptoms in emergency room patients: Results from a two-center randomized controlled study comparing an early single-session Eye Movement Desensitization and Reprocessing intervention with usual care

**DOI:** 10.1101/2023.05.11.23289838

**Authors:** Cédric Gil-Jardiné, Samantha Al Joboory, Juliane Tortes Saint Jammes, Guillaume Durand, Romain Brunschwig, Pierre Catoire, Éric Tellier, Régis Ribéreau-Gayon, Michel Galinski, Louis-Rachid Salmi, Philippe Revel, Guillaume Valdenaire, Karim Tazarourte, Emmanuel Lagarde

**Affiliations:** University Hospital of Bordeaux, Pole of Emergency Medicine, Bordeaux, France; Bordeaux Population Health research center INSERM U1219-”Assessing Health in a Digitalizing real world-setting” team, Bordeaux Cedex, France, E.U.; CASPERTT, Hospital Center of Cadillac, Lormont, France, E.U.; University Hospital Edouard Herriot, Hospices civils de Lyon, Department of Emergency Medicine, Lyon; University Hospital, Claude Bernard University, Lyon, France, E.U.; University Hospital of Bordeaux, Pole of Medicine, Bordeaux, France; EA 7425 Hesper University Hospital, Claude Bernard University, Lyon, France, E.U.

**Author notes:** Corresponding author: Cédric Gil-Jardiné, University Hospital of Bordeaux, Pellegrin Hospital, Emergency Department, Place Amélie, Raba-Léon. 33000 Bordeaux, France, E.U., Mail.

**Keywords:** Stress, emergency room, Eye Movement Desensitization and Reprocessing, post-concussion-like symptoms, post-traumatic stress disorder, clinical trial

## Abstract

**Importance:** After a traumatic event, 10–20% of injured patients will suffer for several months from various symptoms, collectively termed post-concussion-like symptoms (PCLS), which can lead to a decline in quality of life. Moreover, recent findings suggested that this condition may also apply to patients with an acute medical condition. A preliminary randomized controlled trial suggested that this condition may be prevented by a single early short Eye Movement Desensitization and Reprocessing (EMDR) psychotherapeutic session delivered at the ER.

**Objective:** The present study was designed to compare the impact of the early EMDR intervention versus usual care on 3-month PCLS in patients presenting at the ER. Design, Setting, and Participants: This study was an open-label two-center comparative randomized controlled trial with phone follow-up assessments at 3 months. Eligible participants included adults (≥18 years old) presenting at the ER who have a high risk of PCLS using a 3-item scoring scale.

**Interventions:** The randomization groups were as follows: (i) EMDR Recent Traumatic Episode Protocol (R-TEP) intervention performed during the ER stay and (ii) usual care. Main Outcomes and Measures: The primary and secondary outcomes were respectively the frequency of PCLS and PTSD at 3 months after the ER visit.

**Results:** This study included 313 patients with a high risk of PCLS who were randomized into two groups; of these patients, 219 were contacted by phone at 3 months. There was no difference in the primary outcome (EMDR: 53.8% vs. Control: 49.6%), but for the secondary outcome, the occurrence of PTSD was greater in the intervention group (9.4% vs. 2.7%, p = 0.04). In the EMDR group, a high level of self-assessed stress at admission (>6) was strongly associated with persistent PCLS (76.9% vs. 40.9%, p = 0.04).

**Conclusion and Relevance:** The present results showed that a single EMDR R-TEP session did not reduce the incidence of PCLS at 3 months in patients admitted to the ER. However, the rate of PTSD was higher in the EMDR group. These results suggest that more data should be collected to define which treatment options may be offered to patients attending the ER and the role that psychologist skill plays in this process.

**Trial registration:** ClinicalTrials.gov identifier NCT03400813.

**Key points:** *Question:* Does early EMDR in the emergency room reduce the incidence of PCLS at 3 months after care?

*Findings:* In patients admitted to the ER, a single EMDR R-TEP session did not reduce the incidence of PCLS at 3 months, especially among patients who reported a high level of stress at admission.

*Meaning:* The present results suggest that more data will be necessary to determine the available treatment options for patients attending the ER and the role that psychologist skill plays in this process.

## Background

In 2012, the most recent national survey in France revealed that 10.6 million people came or were taken to the emergency room (ER), sometimes on several occasions, as 18 million visits were recorded that year. Although more than 80% of individuals attending the ER leave within a few hours without hospitalization,^1,2^ recent studies^3–6^ have consistently documented that 10–20% of injured patients will suffer for several months from very diverse symptoms after the event and that this may lead to a potentially significant decline in their quality of life. This decline could delay or prevent the resumption of school or work activities and also change social and family relationships. Each year in France, approximately 2 million people are confronted by difficulties of varying degrees, but the causes are often unidentified and may be unrelated to the traumatic event. This relationship remains difficult to understand because these symptoms, including headaches, concentration disorders, memory problems, stress intolerance, personality change, and irritability, appear to be non-specific. Such symptoms have been described for more than 50 years in association with head trauma, and in this context, are referred to as post-concussion syndrome (PCS). However, it is now accepted that these symptoms are not specific to head injuries and can also occur in other patients who visit the ER,^5,7,8^ which greatly expands the size of the affected population. In a cross-sectional observational study of 31,958 high school athletes, Iverson et al.^9^ found that 19% of uninjured boys and 28% of uninjured girls reported having a symptom burden that resembled a diagnosis of PCS based on the International Classification of Diseases, 10th Revision (ICD-10);^9^ subsequently, this diagnosis has frequently been described as post-concussion-like symptoms (PCLS).

The symptoms of PCLS are very similar to, and sometimes exactly the same as, two previously published dimensions of post-traumatic stress disorder (PTSD), i.e., hyperactivation of the nervous system and cognitive and emotional numbing. Thus, most researchers have hypothesized that PCLS and PTSD share, at least in part, the same causal pathway in which stress plays a key role.^8,10,11^ This would be particularly relevant for prevention because only studies that have specifically investigated PTSD are sufficient in number and quality to identify credible modes of intervention.^12^ This led our research group to consider using stress management interventions in the ER in the hope of improving outcomes for traumatized patients. To date, the psychotherapeutic intervention that has proven superior to all other methods for the prevention and treatment of PTSD is Eye Movement Desensitization and Reprocessing (EMDR).^13–17^ In particular, a brief single trauma-focused EMDR protocol, the Recent Traumatic Episode Protocol (R-TEP) method,^18^ was developed and can be used in the context of the ER. EMDR is based on alternated bilateral stimulation that could be conducted using visual (eyes movements) or sensitive (taping) stimulation.

Our research group tested this method in a randomized open-label single-center pilot study of 130 patients with a high risk of PCLS that was conducted in the ER of Bordeaux University Hospital. The patients were randomized into three groups: a 15-minute reassurance session, a 60-minute session of EMDR, and usual care. The proportions of patients with PCLS at 3 months were 18%, 37%, and 65% in the EMDR, reassurance, and control groups, respectively.^19^ The present study was designed to replicate this trial with greater statistical power using patients from two sites.

## Methods

### Study Design

The study population and design of the SymptOms Following Trauma Emergency Response 3 (SOFTER 3) trial have been previously published.^20^ Briefly, this was a two-center open-label randomized controlled trial designed to assess the effects of an early EMDR R-TEP session on PCLS at 3 months compared with those of usual care in patients who presented to the ER. The secondary objectives included comparisons between the EMDR R-TEP and control groups regarding PTSD at 3 months, self-reported stress at ER discharge, self-assessed recovery expectations at discharge and 3 months, and self-reported pain levels at discharge and 3 months.

### Sites and Patients

All patients who came or were brought to the adult ER at one of the study sites following an event that led to an injury or with a new acute medical condition were included in the present study. The inclusion criteria were as follows: ≥18 years of age, conscious, able to provide informed consent, affiliated with social security, and able to understand the study procedures and to comply with them for the entire length of the study; only French speakers were enrolled in the study. Whatever the cause of injury, the event must have occurred in the past 24 hours. Patients who attended the ER for medical reasons were eligible if their condition was acute and if they were presenting to the ER for this reason for the first time. To select the patient most at risk for PCLS at 3 months we used a risk score derived from the results of our previous studies that assessed the determinant of PCLS. This score was computed as follows: female gender, +1; current use of anxiolytics/antidepressant, +1; and perceived health status prior to admission: excellent or very good (0), good (+1), poor (+2), and bad (+3). To be eligible for enrollment in the study, a patient needed to score above the pre-defined threshold of 1 on the 3-item assessment procedure; this was designed to select patients at risk for PCLS. This score was developed using data from previous studies conducted in the ER setting of the Bordeaux Teaching Hospital.^8,10,19^ Patients who were unable to provide written informed consent, unwilling to be contacted at 3 months, and/or under the influence of acute drug or alcohol use or dependence that, in the opinion of the site investigator, could interfere with adherence to study requirements were excluded from the study.

Participants were recruited from among patients who presented to the ERs of the University Hospitals of Bordeaux (Groupe Hospitalier Pellegrin) and Lyon (Groupement Hospitalier Edouard Herriot) and who were determined to have a high risk of PCLS. The identification and recruitment of potential study participants was carried out by emergency personnel under supervision of the project manager. Priority was given to the clinical evaluation and care of each patient, and the recruitment procedure was only initiated when the patient’s condition allowed it. First, oral consent for participation was sought during the risk assessment stage. Then, patients who fulfilled the inclusion criteria and were assessed as having a high risk for PCLS were presented with the objectives and procedures of the study and invited to sign an informed consent form.

#### Data collected

For all patients, we recorded demographic information and the reason for their visit to the ED. Patients were also asked to report their level of stress, recovery expectation and acute pain using a numeric ten-level Likert scale.

### Intervention

At both sites, patients were allocated to one of the two arms of the study using block randomization. Patients in the EMDR group received a 1-hour psychotherapeutic intervention based on the R-TEP protocol,^21^ which incorporates and extends Shapiro’s early EMDR intervention protocols^13^ into an integrative and comprehensive intervention that accounts for the fragmented and unconsolidated nature of recent traumatic memories as well as the need for safety and containment; these sessions were carried out by trained psychologists. Following the eight phases of the standard EMDR protocol it introduces four new procedural concepts (Traumatic Episode, Episode Narrative, “Google Search/ Scan” for identifying disturbing fragments and Current Trauma Focused processing strategies).

All psychologists involved in the study received a one-day specific training for the R-TEP protocol together with a continuous remote mentoring. A standardized questionnaire was completed by the psychologists at the beginning and end of the EMDR session to record the level of disturbance using a Likert scale (0–10) on the Subjective Unit of Disturbance (SUD) scale,^22,23^ and free text commentary was provided to record the details of the session.

In a post-hoc analysis, the skill level of each psychologist was evaluated by an EMDR supervisor blind to the intervention content as well as the 3-month outcomes. Skill level was defined based on professional background, level of formation in EMDR practice (1 or 2), EMDR certification, and experience in the R-TEP protocol prior to the training delivered for the purpose of the study. Fidelity to the protocol was not assessed.

Patients in the usual care group were medically and psychologically managed by the ER staff without the intervention of a study psychologist. Inclusion in the study was only possible on days when psychologists were deployed in the ER.

### Follow-up Assessments

Patients were contacted by phone 3 months after their ER visit using the phone number provided at the time of ER recruitment. Although several attempts were made to contact patients when necessary, the attempts were stopped when the delay exceeded 4 months after ER discharge. Symptoms were assessed using a standardized questionnaire administered by a research assistant blind to the randomization group.

### Outcomes

The primary outcome was the proportion of patients with PCLS at 3 months as measured using the Rivermead Post-Concussion Symptoms Questionnaire.^24^ The definition of PCS in the Rivermead Questionnaire includes the following symptoms: headache, feelings of dizziness, nausea/vomiting, sleep disturbances, fatigue, irritability, noise sensitivity, depression, frustration, poor memory, poor concentration, taking longer to think, blurred vision, light sensitivity, double vision, and restlessness. All variables were measured using a Likert scale that ranged from 0 (not experienced at all) to 4 (a severe problem). Consistent with the PCS definition in the context of mild head injury, patients were defined as having PCLS if they reported at least three symptoms of moderate to high severity.

The secondary outcomes included the presence of PTSD (defined using the PTSD Checklist, 5^th^ version),^25^ self-assessed recovery expectation at discharge, self-reported chronic pain at 3 months, and self-reported acute pain at discharge. All variables were assessed during the 3-month follow-up phone interview.

### Sample Size and Statistical Analysis

Based on previous pilot studies,^19^ this protocol shows a PCLS incidence of 47% in patients with a score ≥2. The goal of the present study was to document a decrease of 15% in PCLS prevalence in the EMDR group. Thus, based on a 5% type I error rate and a power level of 80%, the required sample size was 169 patients in each group. Further considerations for 20% of participants lost to follow-up and 5% lost due to missing data for the main variables resulted in an expected number of 223 patients in each group.

The analyses for the primary and secondary outcomes were conducted on all patients who completed follow-up at 3 months. The prespecified stratified analysis was carried out with considerations for study center, stress level, and individual PCLS risk score. An additional post hoc analysis was conducted in the intervention group to assess the potential impact of psychologist skill level. Differences between patients who completed the study and those who were lost to follow-up were assessed for all variables. All statistical analyses were performed blind to arm allocation.

### Ethics, Confidentiality of Data, and Data and Safety Monitoring Board guidelines

This research project received a positive endorsement from the French Comité de Protection de Personnes (CPP), Ouest II–Angers-N° RCB = 2017-A01462-51–N° CPP = 2017/36. The study was registered on ClinicalTrial.gov (NCT03400813).

## Results

**Table 1.** Patient characteristics

**Table 1:** Patient characteristics.

|  |  | Population Randomized |  |  |  | Completed follow-up |  |  |  |
| --- | --- | --- | --- | --- | --- | --- | --- | --- | --- |
|  |  | EMDR |  | Control |  | EMDR |  | Control |  |
|  |  | n | % | n | % | n | % | n | % |
|  |  | 156 |  | 157 |  | 106 |  | 113 |  |
| **Patients characteristics** |  |  |  |  |  |  |  |  |  |
| Gender | Female | 121 | 77.5 | 114 | 72.6 | 81 | 76.4 | 82 | 72.6 |
| Age * |  | 45 [29-60] |  | 46 [30-62] |  | 50.0 [31-65] |  | 46.0 [30-60] |  |
| Inclusion score | = 2 | 69 | 44.2 | 78 | 49.7 | 51 | 48.1 | 54 | 47.7 |
|  | ≥ 3 | 87 | 55.8 | 79 | 50.3 | 55 | 51.9 | 59 | 52.2 |
| Presence of PCLS at admission |  | 105 | 67.3 | 98 | 62.4 | 70 | 66.0 | 72 | 63.7 |
| Medical disease |  | 55 | 35.3 | 57 | 36.3 | 43 | 40.6 | 44 | 38.9 |
| Trauma condition |  | 67 | 42.9 | 69 | 43.9 | 38 | 35.8 | 47 | 41.6 |
| First ED consultation |  | 142 | 94.0 | 139 | 90.3 | 96 | 92.3 | 103 | 92.0 |
| Tobacco consumption |  | 42 | 27.8 | 51 | 33.6 | 23 | 22.1 | 39 | 35.5 |
| Alcohol consumption |  | 92 | 60.9 | 95 | 61.7 | 63 | 60.6 | 72 | 64.4 |
| Cannabis consumption |  | 18 | 12.1 | 17 | 11.0 | 10 | 9.8 | 12 | 10.7 |
| **At ED admission** |  |  |  |  |  |  |  |  |  |
| Reported pain |  | 117 | 77.5 | 116 | 75.3 | 76 | 73.1 | 82 | 73.2 |
| Self-assessed stress * |  | 4.0 [1.0-6.0] |  | 3.0 [0.0-5.8] |  | 4.5 [1.0-6.3] |  | 3.0 [0.0-6.0] |  |
| Expectation for recovery * |  | 9.0 [7.0-10.0] |  | 9.0 [6.0-10.0] |  | 9.0 [7.5-10.0] |  | 8.0 [5.5-10.0] |  |
| **ED evaluation** |  |  |  |  |  |  |  |  |  |
| Chronic pain reported |  | 79 | 53.7 | 69 | 46.3 | 55 | 54.5 | 53 | 48.6 |
| Chronic pain followed |  | 54 | 66.7 | 47 | 65.3 | 37 | 66.1 | 35 | 62.5 |
| Current daily pain |  | 52 | 41.6 | 64 | 54.7 | 41 | 47.1 | 47 | 51.6 |
| Thinks having been evaluated by psychologist in the ED |  | 154 | 98.7 | 13 | 8.3 | 105 | 99.0 | 12 | 10.6 |
| **At ED discharge** |  |  |  |  |  |  |  |  |  |
| Reported pain |  | 72 | 61.5 | 72 | 63.1 | 51 | 61.4 | 50 | 61.0 |
| Self-assessed stress * |  | 2.0 [0.0-5.0] |  | 1.0 [0.0-5.0] |  | 2.0 [0.0-5.0] |  | 2.0 [0.0-5.0] |  |
| Expectation for recovery * |  | 9.0 [7.0-10.0] |  | 9.0 [5.25-10.0] |  | 9.0 [7.0-10.0] |  | 9.0 [6.0-10.0] |  |
| Satisfaction for ED cares * |  | 9.0 [7.0-10.0] |  | 8.0 [6.0-9.0] |  | 9.0 [7.0-10.0] |  | 8.0 [7.0-9.0] |  |

\* median [interquartile range];
Risk Score = Risk assessment score : Female gender +1; taking at least one anxiolytic treatment +1; Perceived health status priori to admission Excellent. very good 0 . Good : +1. Poor : +2. Bad +3 ; PCLS : Post-concussion like symptoms ; Reported pain : “Do you have pain ?” Yes/No ; Self-assessed stress : 10-level lickert scale from 0 (no pain) to 10 (worst pain imaginable) ; Expectation for recovery: 10-level lickert scale from 0 (no recovery) to 10 (full recovery)

Between January and July of 2018, 1,855 patients were admitted to the ER at times when psychologists were available; of these patients, 313 (200 at Bordeaux and 113 at Lyon) were eligible for the study and were randomized into one of the two groups (156 in the intervention group and 157 in the control group). Of these 313 patients, 94 were lost to follow-up; thus, 219 patients were ultimately included in the final analysis (Fig. 1). Independent of follow-up, the patient characteristics at inclusion were similar between the intervention and control groups (Table 1). The proportion of patients lost to follow-up in the two groups did not differ.

**Figure 1.**
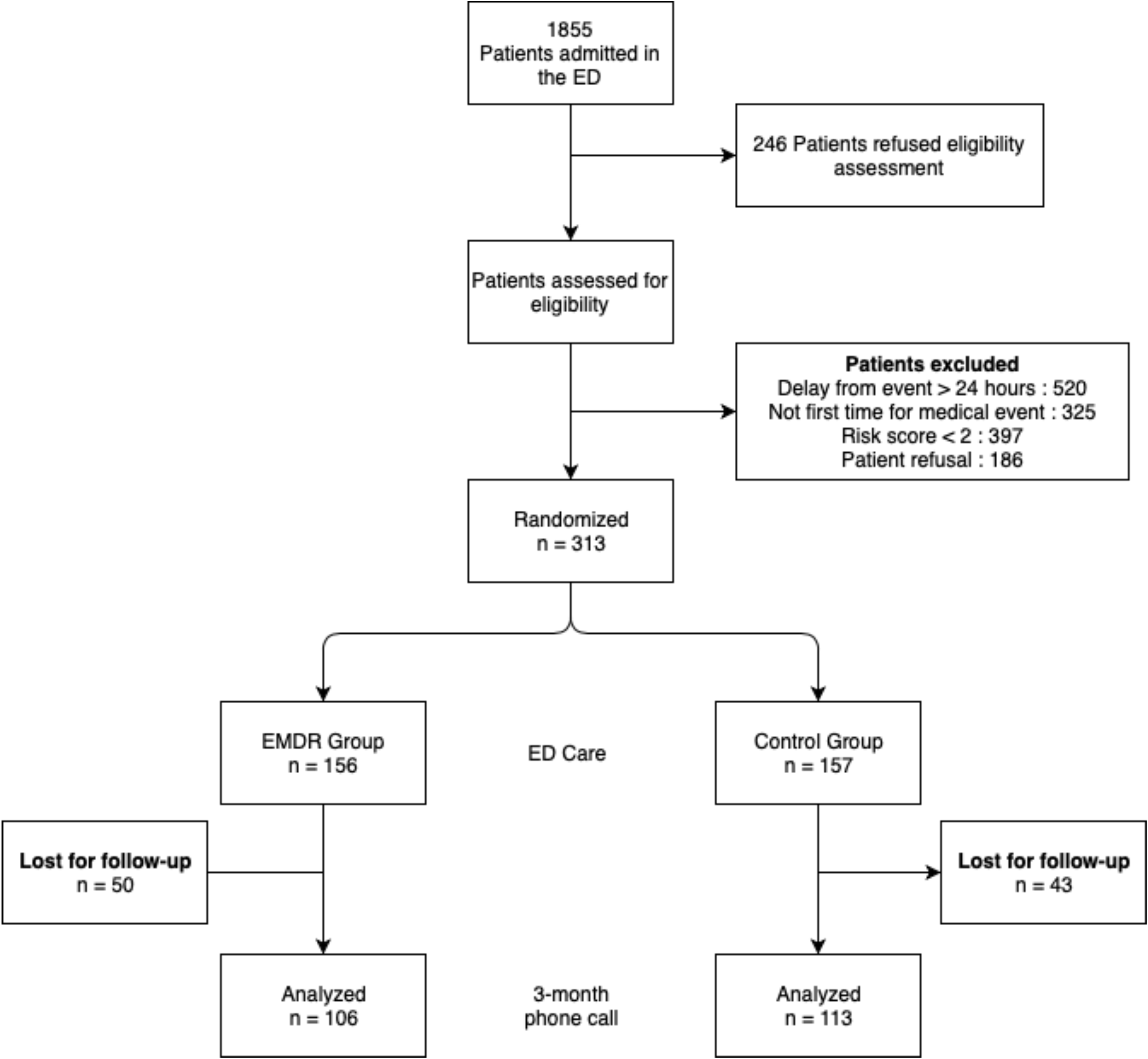
Study Flow Chart.

### Delivery of the Intervention

A total of 31 psychologists participated in the study, representing a total of 984 hours of time present in the ER. All of the psychologists had been previously trained in EMDR^26^ (Level 1: 9; Level 2: 22), 8 had practiced the R-TEP protocol prior to the training delivered for the present study, and 4 were certified in EMDR practice. The median number of interventions performed by each psychologist was three (inter-quartile range: 1.75–4.5). Of the 106 EMDR sessions performed for patients who completed the follow-up assessment, 66 were completed. The median duration of the EMDR sessions was 50 minutes (interquartile range: 30–90); we did not observe any difference according to whether or not a PCLS was present at 3 months. SUD scores decreased between the beginning and end of the EMDR sessions (difference: - 3.9, 95% confidence interval [IC95%]: -4.5 to -3.3).

### Effectiveness

There was no difference between the groups in terms of the primary outcome, i.e., the rate of PCLS (EMDR: 53.8% vs. Control: 49.6%). However, among the secondary outcomes, more cases of PTSD were observed in the intervention group than the control group (9.4% vs. 2.7%, p = 0.04). The occurrence of chronic pain was similar between the two groups (41% vs. 39%, p = 0.78), and the levels of acute pain at discharge did not differ (median [inter-quartile range]: 9 [7–10] vs. 9 [6–10], p = 0.89).

**Table 2.** Primary and secondary outcomes.

**Table 2:** Presentation of the impact of psychologist skill level on PCLS occurrence at 3 months.

|  | Population |  | PCS + |  | PCS- |  |
| --- | --- | --- | --- | --- | --- | --- |
|  | N | % | n | % | n | % |
| **Level of EMDR training** |  |  |  |  |  |  |
| **N1** | 34 | 33.3 | 17 | 89.5 | 17 | 85.0 |
| **N2** | 68 | 66.7 | 2 | 10.5 | 3 | 15.0 |
| **Certification** |  |  |  |  |  |  |
| **Yes** | 7 | 6.9 | 3 | 5.5 | 4 | 8.5 |
| **No** | 95 | 93.1 | 52 | 94.5 | 43 | 91.5 |
| **Experienced in R-TEP EMDR practice before study** |  |  |  |  |  |  |
| **Yes** | 15 | 14.7 | 6 | 10.9 | 9 | 19.1 |
| **No** | 87 | 85.3 | 49 | 89.1 | 38 | 80.9 |
| **Psychologist skill level** |  |  |  |  |  |  |
| **A** | 13 | 12.7 | 5 | 9.1 | 8 | 17.0 |
| **B** | 32 | 31.4 | 16 | 29.1 | 16 | 34.0 |
| **C** | 53 | 52.0 | 31 | 56.4 | 22 | 46.8 |
| **D** | 4 | 3.9 | 3 | 5.4 | 1 | 2.1 |

Skill level of the psychologist was evaluated by an EMDR supervisor blinded from both interventions delivery and 3-month outcomes

**Table 3.** Presentation of the impact of psychologist skill level on PCLS occurrence at 3 months.

**Table 3:** Primary and secondary outcomes.

| **Variable** | **Population** | **EMDR** | **Control** | **p-value** |
| --- | --- | --- | --- | --- |
|  | **N** | **N** | **N** |  |
|  | % [CI 95%] | % [CI 95%] | % [CI 95%] |  |
| **Primary outcome** |  |  |  |  |
| **Number of patients** | 219 | 106 | 113 |  |
| **PCLS** | 53.5%<br>[43.9 to 63.4] | 53.8%<br>[43.9% to 63.4%] | 49.6%<br>[40.1% to 59.1%] | 0.58 |
| **Secondary outcomes** |  |  |  |  |
| **Number of patients** | 219 | 106 | 113 |  |
| **PTSD** | 5.9%<br>[3.3% to 10.2%] | 9.4<br>[4.8% to 17.1%] | 2.7%<br>[0.7% to 8.1%] | 0.04 |
| **Number of patients** | 165 | 83 | 82 |  |
| **Acute pain at discharge** | 61.2%<br>[53.3% to 68.6%] | 61.4%<br>[50.1% to 71.7%] | 61.0%<br>[49.5% to 71.4%] | 1 |
| **Number of patients** | 218 | 106 | 112 |  |
| **Chronic pain at 3-months** | 39.4%<br>[33.0% to 46.3%] | 40.6%<br>[31.3% to 50.6%] | 38.6%<br>[29.5% to 48.1%] | 0.78 |
| **Number of patient** | 162 | 80 | 82 |  |
| **Expectation for recovery\*** | 9 [6 – 10] | 9 [7 – 10] | 9 [6 – 10] | 0.89 |

PCLS: Post-concussion like symptoms
PTSD: Post traumatic stress disorder
\*median [inter-quartile range]

### Post hoc Analyses

The analysis of PCLS according to psychologist skill level indicated that the qualifications of the practitioner may have influenced the outcome because the incidence of PCLS at 3 months was lower among patients who were seen by the most qualified and skilled psychologists (Table 2). There was no association between an incomplete session and an increased risk of PCLS. However, a high self-assessed stress level at admission (>6) was strongly associated with an increased risk of PCLS in the EMDR group (76.9% vs. 40.9%; Table 3 and Fig. 2).

**Figure 2.**
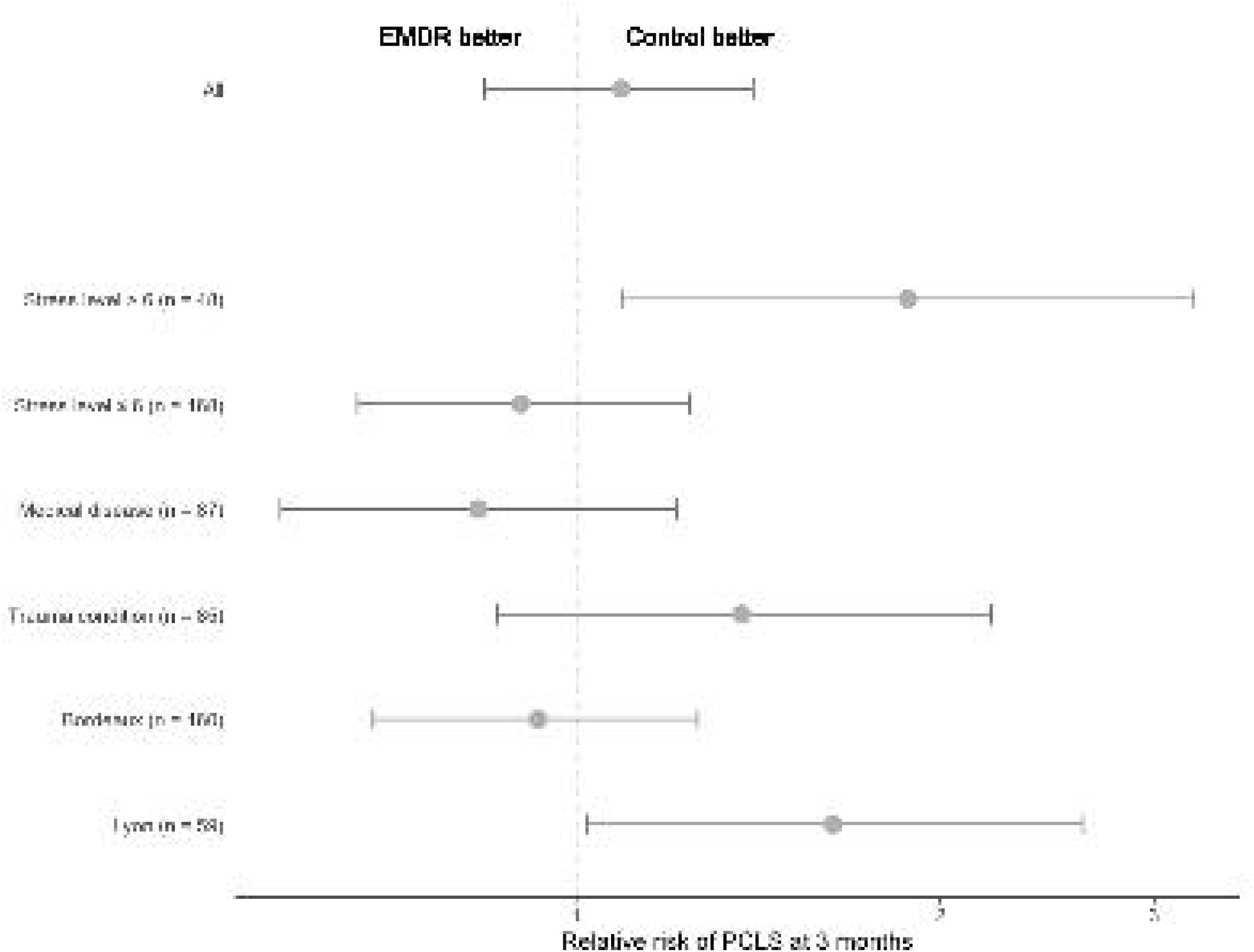
Subgroup analysis: Relative Risk of post-concussion like symptoms occurrence after stratification on different factors.

The overall incidence of PCLS did not differ between the two study centers (Bordeaux: 50.7%, IC95%: 41.4–57.4; Lyon: 54.2%, IC95%: 32.9–59.2). However, the incidence of PCLS in the EMDR group was 48.8% (IC95%: 37.5–60.1) at Bordeaux and 69.2% (IC95%: 48.1–84.9) at Lyon. Figure 2 presented Relatives Risks of post-concussion like symptoms occurrence after stratification on different factors. The difference in PCLS incidence between the intervention and control groups according was not related to patients’ reasons for attending the ER.

## Discussion

The results of the present trial revealed that an early EMDR R-TEP session performed during the ER stay did not reduce the incidence of PCLS at 3 months compared to usual ER care. Moreover, there was a higher incidence of PTSD in the intervention group, and the intervention resulted in an increased incidence of PCLS at 3 months among patients in the highest quartile of self-assessed stress at admission. Finally, there was an association between psychologist qualification level and success of the intervention.

The present study failed to confirm the results obtained during the SOFTER 2 trial.^19^ In that study, there was a substantially lower rate of PCLS among patients treated by a psychologist in an EMDR session compared to those treated with usual care in the ER. More specifically, 6 of 34 patients in the EMDR group had PCLS at 3 months compared with 24 of 37 patients in the control group. There are several possible reasons for this discrepancy between the studies. Only two experienced psychologists were involved in the previous pilot study, whereas 31 psychologists with heterogeneous levels of experience were recruited for the present trial. Of these 31 psychologists, only 8 had previous experience with the R-TEP protocol. The present study found clear positive associations between the outcome of the intervention and the various indicators used to assess the psychologists’ experience and skill. Although it is possible that this can explain the present results, the assessment of the psychologists’ competencies was not planned in the initial protocol and was only conducted after the effectiveness results were known. Therefore, this should remain a hypothesis, but it is also indicative of the need to carefully control the level of training provided to EMDR therapists because the short training period may have been insufficient.^27^ Having less experienced and/or trained psychologists might also have reduced patient adherence to the protocol and increased the number of refusals. Thus, future studies should evaluate fidelity to the intervention protocol.

Approximately 30% of patients included in the present trial were lost to follow-up, but the characteristics of the patients who answered the 3-month questionnaire did not differ from those who did not. The proportion of refusals in the SOFTER 3 trial (∼40%) was significantly higher than that in the SOFTER 2 trial (∼20%). There is no clear explanation for this difference, and it may have influenced the results. In fact, it is possible that the patients who agreed to participate in the study differed from those who did not, which might explain why the expected number of patients was not achieved.

Further analysis of the discrepancies between the present trial and the previous pilot study, which produced more encouraging results, revealed differences in the psychologists’ reports about the nature of the points of disturbance in the EMDR sessions. In the pilot study, the psychologists primarily addressed issues that were not directly related to the event that led a patient to the ER, whereas in the present study, a majority of the intervention reports mentioned disturbance points that were directly related to the event. It was also noted that patients with 3-month PCLS exhibited a significant decrease in SUD scores between the beginning and end of the EMDR session.

The present findings also differed from those of some studies in the literature.^17,19,28^ A study conducted in Israel reported very promising results following a single session of early modified EMDR provided in a general hospital setting by psychologists who were experienced in EMDR practice.^28^ In that study, patients reported the presence of acute stress syndrome and suffered from intrusion distress following accidents and terrorist bombing attacks. However, at the 4-week and 6-month follow-up assessments, the immediate responders in the terror victims group remained symptom free.

The second key finding of the present study concerned the high level of adverse effects associated with the intervention in patients who described themselves as experiencing high levels of stress. When EMDR is performed by an unqualified practitioner, insufficient attention may be paid to the importance of initially establishing sufficient stabilization and calming, which should be part of the protocol when applied correctly. Importantly, the issue of managing patients with high levels of stress or dissociation remains. In response to this challenge, modified and adapted EMDR-type early intervention protocols have been developed to assist victims.^21,29^

Additionally, in the present study, perceived stress was evaluated using a 10-point Likert scale that had never been validated for stress assessment in the ER. Nonetheless, this scale provided a method with which to measure variations in subjective stress between admission and discharge, and a similar 5-point Likert scale for acute stress (“not at all” to “strongly”) had previously been validated.^30^ The use of a 10-point scale likely did not influence the validity of the acute stress measure, especially because this variable was a secondary outcome and was assessed in a post hoc analysis. However, patients in the EMDR group who were experiencing high stress, defined as a numeric score >6, reported many more symptoms than did those in the control group.

## Conclusions

Among patients admitted to the ER in the present study, a single EMDR R-TEP session did not reduce the incidence of PCLS at 3 months, particularly among patients who reported high levels of stress at admission. The present results suggest that it will be necessary to collect more data to determine the available treatment options that can be offered to patients attending the ER. Furthermore, the present results must be applied with caution, particularly due to the large degree of heterogeneity in the skill level of the psychologists employed in this study. Regardless of this issue, clinicians should continue attempts to identify the best care options for traumatized patients who present to the ER.

## Data Availability

All data produced in the present study are available upon reasonable request to the authors

## Data Availability

All data produced in the present study are available upon reasonable request to the authors

## References

1. Carrasco V, Baubeau D. Les usagers des urgences. Premiers résultats d’une enquête nationale. Etudes et résultats. Published online 2003:8.

2. Vuagnat A. Les Urgences Hospitalières, Qu’en Sait-On. Le Panorama des établissements de santé–édition,; 013.

3. de Leon MB, Kirsch NL, Maio RF, et al. Baseline predictors of fatigue 1 year after mild head injury. Arch Phys Med Rehabil. 2009;90(6):956–965. doi:10.1016/j.apmr.2008.12.016

4. Friedland JF, Dawson DR. Function after motor vehicle accidents: a prospective study of mild head injury and posttraumatic stress. J Nerv Ment Dis. 2001;189(7):426–434.

5. McLean SA, Kirsch NL, Tan-Schriner CU, et al. Health status, not head injury, predicts concussion symptoms after minor injury. Am J Emerg Med. 2009;27(2):182–190. doi:10.1016/j.ajem.2008.01.054

6. Stovner LJ, Schrader H, Mickeviciene D, Surkiene D, Sand T. Headache after concussion. Eur J Neurol. 2009;16(1):112–120. doi:10.1111/j.1468-1331.2008.02363.x

7. Smith-Seemiller L, Fow NR, Kant R, Franzen MD. Presence of post-concussion syndrome symptoms in patients with chronic pain vs mild traumatic brain injury. Brain Inj. 2003;17(3):199–206.

8. Laborey M, Masson F, Ribéreau-Gayon R, Zongo D, Salmi LR, Lagarde E. Specificity of postconcussion symptoms at 3 months after mild traumatic brain injury: results from a comparative cohort study. J Head Trauma Rehabil. 2014;29(1):E28–36. doi:10.1097/HTR.0b013e318280f896

9. Iverson GL, Silverberg ND, Mannix R, et al. Factors Associated With Concussion-like Symptom Reporting in High School Athletes. JAMA Pediatr. 2015;169(12):1132–1140. doi:10.1001/jamapediatrics.2015.2374

10. Lagarde E, Salmi LR, Holm LW, et al. Association of symptoms following mild traumatic brain injury with posttraumatic stress disorder vs. postconcussion syndrome. JAMA Psychiatry. 2014;71(9):1032–1040. doi:10.1001/jamapsychiatry.2014.666

11. Gil-Jardiné C, Hoareau S, Valdenaire G, et al. Stress and lasting symptoms following injury: Results from a 4-month cohort of trauma patients recruited at the emergency department. Int Emerg Nurs. Published online November 2019:100810. doi:10.1016/j.ienj.2019.100810

12. Bryant R. Post-traumatic stress disorder vs traumatic brain injury. Dialogues Clin Neurosci. 2011;13(3):251–262.

13. Shapiro F. Eye Movement Desensitization and Reprocessing (EMDR): Basic Principles, Protocols, and Procedures. 2nd ed. Guilford Press; 2001.

14. Bradley MC, Fahey T, Cahir C, et al. Potentially inappropriate prescribing and cost outcomes for older people: a cross-sectional study using the Northern Ireland Enhanced Prescribing Database. Eur J Clin Pharmacol. 2012;68(10):1425–1433. doi:10.1007/s00228-012-1249-y

15. Seidler GH, Wagner FE. Comparing the efficacy of EMDR and trauma-focused cognitive-behavioral therapy in the treatment of PTSD: a meta-analytic study. Psychol Med. 2006;36(11):1515–1522. doi:10.1017/S0033291706007963

16. Bisson JI, Roberts NP, Andrew M, Cooper R, Lewis C. Psychological therapies for chronic post-traumatic stress disorder (PTSD) in adults. Cochrane Database Syst Rev. 2013;(12):CD003388. doi:10.1002/14651858.CD003388.pub4

17. Tarquinio C, Rotonda C, Houllé WA, et al. Early Psychological Preventive Intervention For Workplace Violence: A Randomized Controlled Explorative and Comparative Study Between EMDR-Recent Event and Critical Incident Stress Debriefing. Issues Ment Health Nurs. 2016;37(11):787–799. doi:10.1080/01612840.2016.1224282

18. Shapiro E, Laub B. Early EMDR Intervention (EEI): A Summary, a Theoretical Model, and the Recent Traumatic Episode Protocol (R-TEP). J EMDR Pract Res. 2008;2(2):79–96. doi:10.1891/1933-3196.2.2.79

19. Gil-Jardiné C, Evrard G, Al Joboory S, et al. Emergency room intervention to prevent post concussion-like symptoms and post-traumatic stress disorder. A pilot randomized controlled study of a brief eye movement desensitization and reprocessing intervention versus reassurance or usual care. J Psychiatr Res. 2018;103:229–236. doi:10.1016/j.jpsychires.2018.05.024

20. Gil-Jardiné C, Al Joboory S, Tortes Saint Jammes J, et al. Prevention of post-concussion-like symptoms in patients presenting at the emergency room, early single eye movement desensitization, and reprocessing intervention versus usual care: study protocol for a two-center randomized controlled trial. Trials. 2018;19(1):555. doi:10.1186/s13063-018-2902-2

21. Shapiro E. EMDR and early psychological intervention following trauma. Rev Eur Psychol AppliquéeEuropean Rev Appl Psychol. 2012;62(4):241–251. doi:10.1016/j.erap.2012.09.003

22. Wolpe J. The Practice of Behavior Therapy. Pergamon Press; 1990.

23. Wolpe J, Abrams J. Post-traumatic stress disorder overcome by eye-movement desensitization: a case report. J Behav Ther Exp Psychiatry. 1991;22(1):39–43.

24. King NS, Crawford S, Wenden FJ, Moss NE, Wade DT. The Rivermead Post Concussion Symptoms Questionnaire: a measure of symptoms commonly experienced after head injury and its reliability. J Neurol. 1995;242(9):587–592.

25. Blanchard EB, Jones-Alexander J, Buckley TC, Forneris CA. Psychometric properties of the PTSD Checklist (PCL). Behav Res Ther. 1996;34(8):669–673.

26. FORMATION EMDR INITIALE. Accessed May 24, 2019. http://www.efpe.fr/formation-initiale.html#formation

27. Chen YR, Hung KW, Tsai JC, et al. Efficacy of Eye-Movement Desensitization and Reprocessing for Patients with Posttraumatic-Stress Disorder: A Meta-Analysis of Randomized Controlled Trials. PLOS ONE. 2014;9(8):e103676. doi:10.1371/journal.pone.0103676

28. Kutz I, Resnik V, Dekel R. The Effect of Single-Session Modified EMDR on Acute Stress Syndromes. J EMDR Pract Res. 2008;2(3):190–200. doi:10.1891/1933-3196.2.3.190

29. Shapiro, E., & Laub, B. The recent traumatic episode protocol (R-TEP): An integrative protocol for early EMDR intervention (EEI). In: Implementing EMDR Early Mental Health Interventions for Man-Made and Natural Disasters: Models, Scripted Protocols and Summary Sheets. 1 edition. Springer Publishing Company; 2013.

30. Hampel P, Petermann F. Perceived stress, coping, and adjustment in adolescents. J Adolesc Health Off Publ Soc Adolesc Med. 2006;38(4):409–415. doi:10.1016/j.jadohealth.2005.02.014

